# Prediction of post-operative delirium with machine learning in abdominal surgery with comorbidity indices and laboratory values

**DOI:** 10.64898/2026.06.28.26356787

**Authors:** Wesley Chorney, Shinhee Kang, Sing Hui Ling, Michael Lisi

## Abstract

**Background:** Postoperative delirium (POD) is a complication associated with most types of surgery, and is associated with a number of detrimental effects. Therefore, it is of interest to determine which patients may be at higher risk of POD so that mitigating steps may be taken. We sought to determine whether POD can be accurately predicted with common machine learning (ML) models.

**Methods:** Using the Medical Information Mart for Intensive Care (MIMIC)-IV database, we identified 8026 abdominal surgery procedures across 7215 adult patients. Using demographic information, such as age, type of surgery, sex; as well as commonly measured laboratory values (such as electrolytes and blood counts) and comorbidity indices, we determined to what extend common ML models, such as random forests, support vector machines, extreme gradient boosted machines, and neural networks, could predict POD.

**Results:** Random forests outperformed logistic regression, support vector machines, extreme gradient boosted machines, and neural networks, with respect to individual *t*-tests. The random forest model had a sensitivity of 73.11, a specificity of 71.14, and an area under the receiver operator characteristic curve of 0.800. Age, comorbidity indices, gender, and alcohol use carried significant predictive weight in this cohort.

**Conclusions:** Machine learning models are effective predictors of postoperative delirium, although further work is required to increase clinical utility of such tools. Markers of inflammation, comorbidity indices, and alcohol use are important predictive features alongside better-known features such as age.

## Introduction

Postoperative delirium (POD) is a significant complication of many types of surgeries. It is an incompletely understood phenomenon, with a number of proposed mechanisms, including the stress response to surgery provoking pathological elevations in cortisol levels [1], imbalance in neurotransmitters such as acetylcholine, *γ*-aminobutyric acid (GABA), norepinephrine, serotonin, and dopamine [2]; neuroinflammation due to proinflammatory cytokines from surgical trauma, anesthesia, and postoperative pain [3], which together with a weakened blood-brain barrier with increasing age [4], may be a significant contributor of the increased incidence of POD in elderly individuals. Additionally, disruption of neural networks in the brain [5] and genetics [6] are contributing factors to POD.

Although the exact pathophysiology remains incompletely understood, POD is an important topic of investigation because it has profound effects for patients, their carers, and the healthcare system in general. For patients, POD is associated with increased postoperative mortality [7] and functional decline [8]; whereas for carers, there is an increased burden of care and risk of caregiver burnout [9]. Finally, POD is associated with increased healthcare burden and costs [10, 11]. Certain factors are known to be associated with POD — perhaps the most well-known is age, which has been corroborated in numerous studies [12–14], but also factors such as Parkinson’s disease [15], depression [16], alcohol use [17, 18], and dementia [19]. Additionally, comorbidity indices, such as the Charlson comorbidity index [20] and the Elixhauser comorbidity index [21], which associate weightings with different (chronic) conditions to derive an overall picture of comorbidity, are associated with POD in abdominal surgery [22]. Additionally, laboratory markers such as elevated platelets and sodium have been shown to be associated with POD [23].

While the above individual risk factors are strongly associated with POD, the actual pattern amongst these and other risk factors and the development of POD is almost certainly far more complex. Therefore, machine learning (ML) models may be of use in modelling the association between preoperative risk factors and POD. Studies have investigated how ML models can aid in the prediction of POD across all types of surgery [24], non-cardiac surgery [25], cardiac surgery [26], and most frequently in the elderly population [27–30]. Between studies, the type of input data varies significantly, including intraoperative electroencephalographs [30], intraoperative blood loss, anesthesia duration, and intubation time [31], comorbidity indices [22], polysomnography [32], and abnormal laboratory results [33].

Despite the wealth of previous studies in the prediction of POD, we note some important caveats. First, either the specific type of surgery should be indicated in the input to an ML model, or models should be trained on data from a specific type of surgery only. This is due to the fact that different types of surgeries induce different levels of stress [34], which is a proposed mechanism of POD. Therefore, some signal is most likely lost when indications of the type of surgery omitted. Second, ideally the population under study should be broadened beyond only elderly patients. While not as common, POD can occur in non-elderly adults [35] and even pediatric patients (though in this specific group, contributing factors likely differ) [36]. Finally, careful attention should be paid to input variables, keeping in mind the purpose of a predictive model. While there is utility in predicting which patients may develop POD based on intraoperative variables, ideally high-risk cases could be flagged prior to surgery, since there is significant burden to the patient. This could allow for better discussions around informed consent and exploration of alternate treatments. To address all of the above, we propose a ML model for the prediction of POD

- in patients undergoing abdominal surgery specifically,
- who are adults of any age,
- and using only preoperative variables, including demographics, comorbidity indices, and preoperative laboratory values.

## Materials and methods

### Dataset

This retrospective observational study utilized data from the Medical Information Mart for Intensive Care IV (MIMIC-IV), which is a publicly available, de-identified electronic health record database comprising admissions to the Beth Israel Deaconess Medical Center in Boston, Massachusetts, between 2008 and 2022 [37]. MIMIC-IV is freely available via PhysioNet [38]. MIMIC-IV was developed in accordance with the Health Insurance Portability and Accountability Act (HIPAA) Safe Harbor provisions, with all protected health information removed or appropriately de-identified before release. The database is maintained under institutional review board (IRB) approval from the Beth Israel Deaconess Medical Center and the Massachusetts Institute of Technology, with a waiver of informed consent due to the retrospective use of de-identified data.

Adult patients undergoing major general surgical procedures were identified using both International Classification of Diseases (ICD)-9 procedure codes and ICD-10 procedure codes corresponding to common general surgical operations, including cholecystectomy, appendectomy, colorectal resection, gastrectomy, bariatric surgery, inguinal, ventral, and incisional hernia repair, adrenalectomy, splenectomy, hepatectomy, pancreatectomy, exploratory laparotomy, and bowel resection. When multiple eligible procedures occurred during a single hospitalization, only the earliest qualifying operation was retained to ensure a single index procedure per patient.

Patient-level demographic and clinical variables extracted for analysis included age at surgery, sex, admission type (elective versus emergency), Charlson Comorbidity Index, and Elixhauser comorbidity score. The Elixhauser score was derived using ICD-10 van Walraven weights [39] when ICD-10 diagnosis codes were available and ICD-9 Quan weights otherwise [40], thereby permitting standardized comorbidity assessment across both coding eras. Additional clinically relevant comorbidities, including dementia, Parkinson disease, depression, and alcohol use disorder, were also identified from diagnosis codes. Preoperative laboratory investigations obtained within the seven days preceding surgery were extracted from the hospital laboratory database. For each laboratory analyte, the minimum and maximum recorded values during the preoperative window were calculated. These minimum and maximum values have been shown to carry important prognostic information across different surgery types [23, 41–43]. Laboratory variables included complete blood count variables (hemoglobin, hematocrit, red blood cell count, white blood cell count, platelet count, and differential cell counts), renal function markers (creatinine, blood urea nitrogen, estimated glomerular filtration rate, sodium, potassium, chloride, bicarbonate, calcium, magnesium, and phosphate), liver function tests (alanine aminotransferase, aspartate aminotransferase, alkaline phosphatase, total bilirubin, albumin, and total protein), inflammatory markers when available, coagulation studies, and thyroid function tests (thyroid-stimulating hormone and free thyroxine).

Overall, there were 8026 abdominal surgery procedures across 7215 patients. In this cohort, there were 92 cases of postoperative delirium (corresponding to an incidence of approximately 1.15%).

### Machine learning models

We trained logistic regression, support vector machines, neural networks, extreme gradient boosted machines (XGBoost), and random forests on the dataset. Both random forests and extreme gradient boosted machines rely in part on ensembling, which is the principle of combining a large number of weak classifiers trained on all or part of the training data to create a stronger classifier through reducing bias while still capturing variance [44]. XGBoost uses gradient boosting to ensure that classifiers added to the ensemble address residual error in each iteration [45]. Support vector machines find a hyperplane that maximizes the separation between each class. This is often facilitated by a kernel-trick [46], where the input data is embedded into a higher dimension by means of multiple transformations to achieve better separation between the positive and negative class of the input data. Neural networks are ubiquitous in applications of ML methods to healthcare, as they can deal with a wide variety of input data, and are able to model patterns of arbitrary complexity. Applications range from analysis of computed tomography imaging [47], to evaluation of surgical skills [48], to analysis of electrocardiograms distributed amongst different institutions [49]. Given the tabular nature of the training data, the neural network we chose to evaluate was a multilayer perceptron with four layers.

### Analysis

We used stratified 10-fold cross validation to evaluate each model. Given the significantly imbalanced nature of the dataset, models were evaluated principally on area under the receiver operator characteristic curve (AUC-ROC). For each model, we also report the accuracy, precision, sensitivity, specificity, F1 score, and the area under the precision-recall curve (AUC-PR). Analysis was carried out using Python 3.13.5 [50]. Comparisons between models were made using *t*-tests with level of significance *α* = 0.05, with the assumption of normality.

### Ethics

#### Human Participants

This study used de-identified data from the MIMIC-IV database, hosted on PhysioNet [38]. The collection of patient information and creation of the MIMIC-IV resource were reviewed and approved by the Institutional Review Boards (IRBs) of the Massachusetts Institute of Technology (MIT, Cambridge, MA, USA) and Beth Israel Deaconess Medical Center (BIDMC, Boston, MA, USA) (MIT IRB #0403000206; BIDMC IRB #2001-P-001699/14), who granted a waiver of informed consent and authorized data sharing.

All patient data are fully de-identified in accordance with the Health Insurance Portability and Accountability Act (HIPAA) Privacy Rule. Therefore, the use of MIMIC-IV data for research purposes is considered exempt from additional institutional review board approval or informed consent requirements.

The required training to access the database was completed by the first author.

#### Participant Consent

This study did not involve direct interaction with human participants. The analysis was performed using the MIMIC-IV database.

The collection of patient data and creation of the MIMIC-IV resource were reviewed and approved by the Institutional Review Boards (IRBs) of the Massachusetts Institute of Technology (MIT, Cambridge, MA, USA) and Beth Israel Deaconess Medical Center (BIDMC, Boston, MA, USA) (MIT IRB #0403000206; BIDMC IRB #2001-P-001699/14). A waiver of informed consent was granted due to the retrospective nature of the study and the use of fully de-identified health records in compliance with the Health Insurance Portability and Accountability Act (HIPAA) Privacy Rule.

As the present study used only de-identified data, no additional participant consent was required.

## Results

Table 1 gives an overview of the main results. For each model, the mean accuracy, precision, sensitivity, specificity, F1 score, AUC-ROC, and AUC-PR across all ten folds are displayed, plus or minus standard deviation. We use bold font to highlight the best metric across all models. Overall, the random forest classifier performs the best, though it is outperformed by the support vector machine with respect to sensitivity and AUC-PR. With respect to AUC-ROC, the random forest classifier outperforms the logistic regression and neural network in a statistically significant manner (*p* = 0.0093, *p* = 0.0002, respectively) but does not outperform the support vector machine or XGBoost classifier with statistical significance (*p* = 0.0786, *p* = 0.1449, respectively). While it has the best AUC-ROC, the relatively lower sensitivity is due to the failure of the random forest classifier to correctly identify true positives.

**Table 1.** Results for each model, represented as mean plus or minus standard deviation. The best metric across all models is shown in bold.

| Model | Accuracy | Precision | Sensitivity | Specificity | F1 Score | AUC-ROC | AUC-PR |
| --- | --- | --- | --- | --- | --- | --- | --- |
| Logistic Regression | 62.51±2.84 | 2.01±0.37 | 66.11±10.83 | 62.47±2.89 | 3.91±0.71 | 0.714±0.061 | 0.049±0.030 |
| Random Forest | **71.16±1.39** | **2.84±0.41** | 73.11±13.14 | **71.14±1.46** | **5.46±0.80** | **0.800±0.044** | 0.053±0.019 |
| Support Vector Machine | 65.65±6.11 | 2.53±0.59 | **75.11±14.50** | 65.54±6.21 | 4.88±1.13 | 0.764±0.056 | **0.065±0.037** |
| XGBoost | 68.38±1.97 | 2.65±0.55 | 74.22±14.79 | 68.31±1.93 | 5.11±1.05 | 0.769±0.062 | 0.053±0.018 |
| Neural Network | 63.64±2.67 | 2.22±0.39 | 71.78±15.13 | 63.55±2.80 | 4.31±0.76 | 0.740±0.051 | 0.041±0.022 |

Fig 1 displays the receiver operator characteristic curve for the random forest model versus a random chance classifier. The initially low slope indicates that as the threshold increases, the false positive rate increases disproportionately to the true positive rate. Nevertheless, the relatively high AUC-ROC (a perfect classifier would have an AUC-ROC of 1.00) indicates good discrimination at different thresholds between classes.

**Fig 1.**
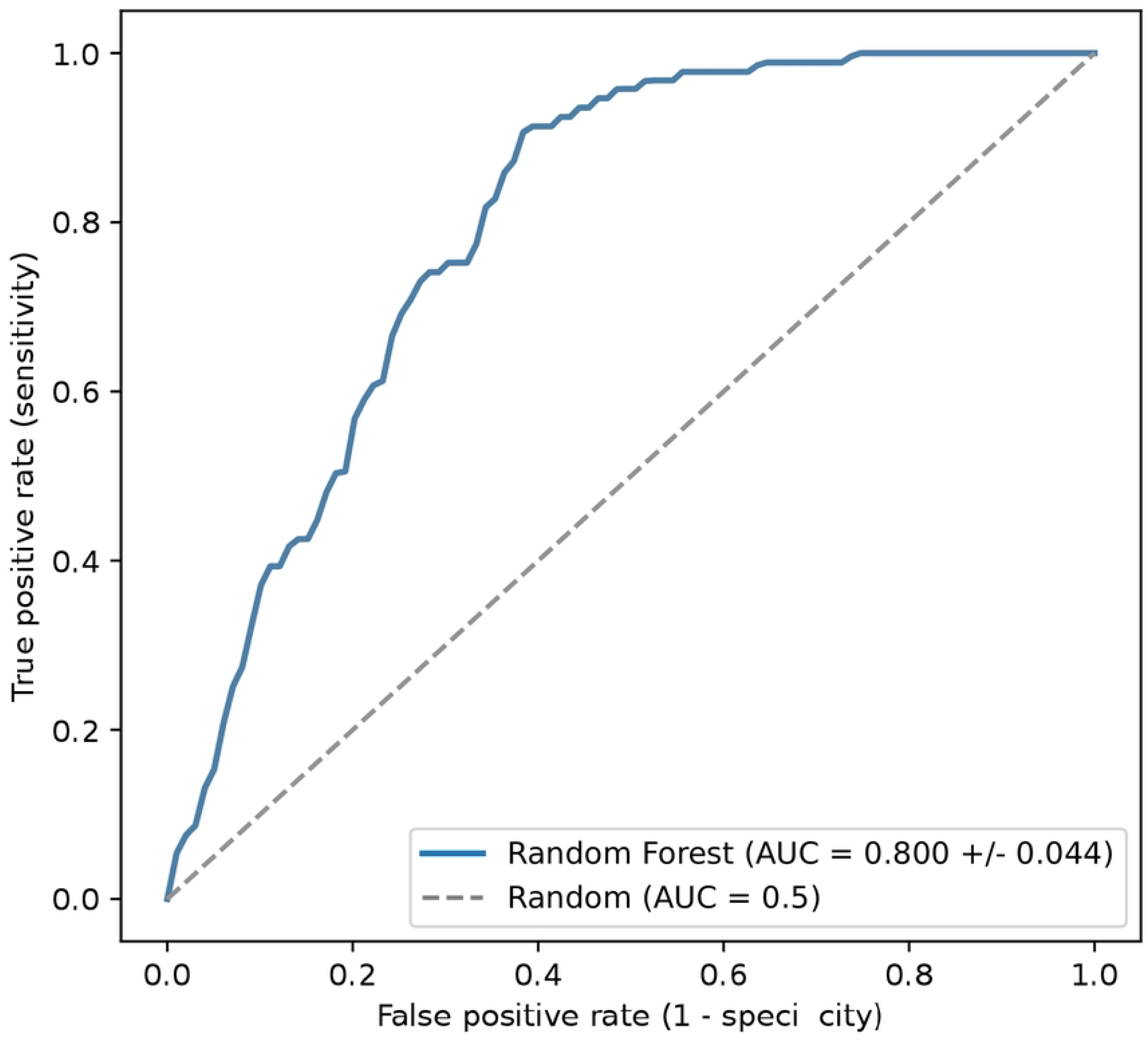
Receiver operator characteristic curve for the random forest model, with area under the curve displayed in the legend, versus a random chance classifier.

Fig 2 displays the predicted probability of delirium per class for the random classifier. We note that for the patients who do not experience postoperative delirium, there is a relatively smooth probability distribution, although with an ideal classifier the distribution would be less right-skewed. The distribution for patients who do experience postoperative delirium is less smooth, most likely due to the relatively low incidence in this population.

**Fig 2.**
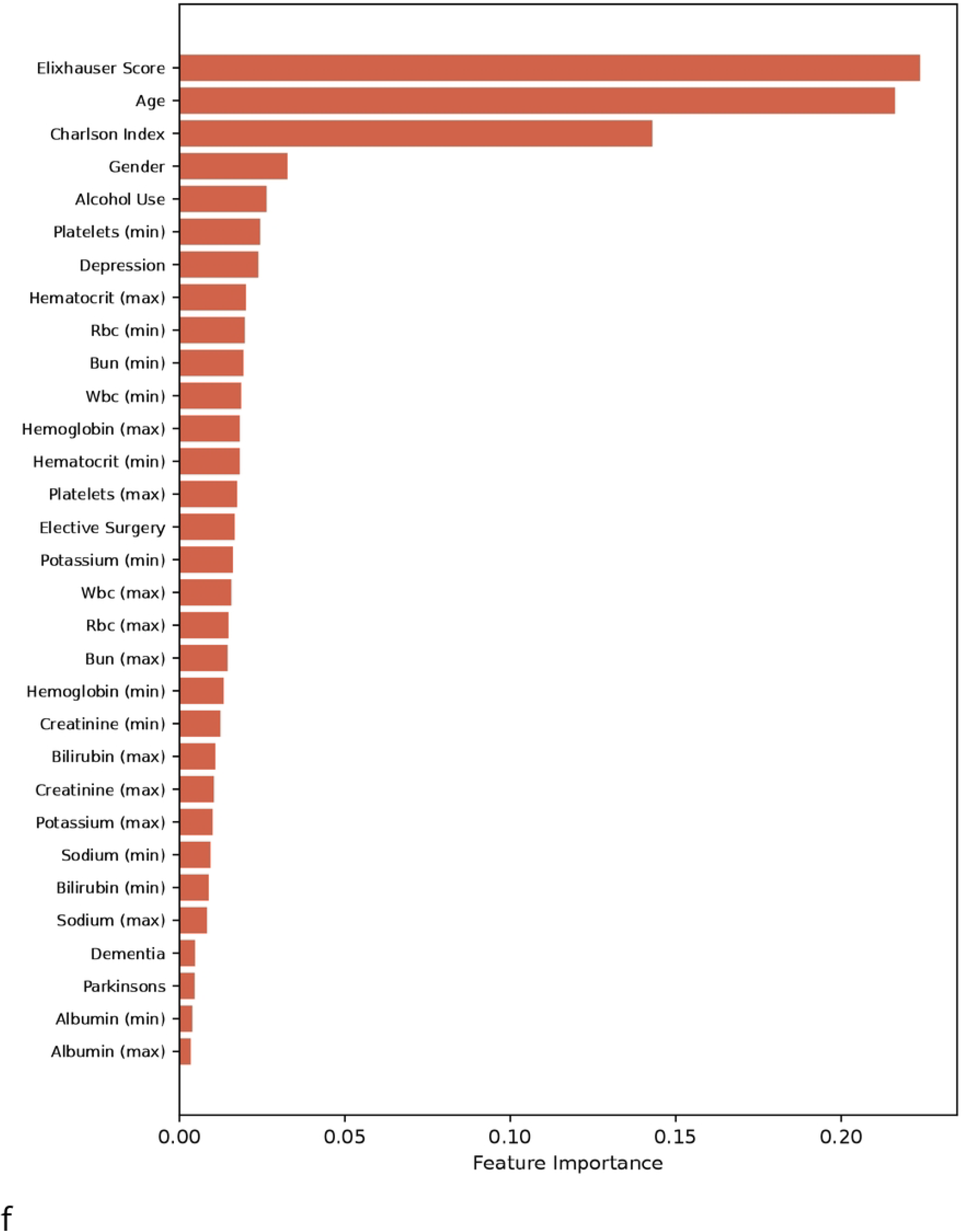
Probability distribution of model predictions versus true class for the random forest classifier.

Finally, Fig 3 shows the feature importance for each input variable to the random forest. These features were obtained via mean decrease in impurity. Ideally, any feature added to a decision tree should help the classifier better segregate positive and negative classes, which leads to a decrease in impurity. By calculating the mean decrease in impurity over all decision trees with said feature, a relative importance of a given feature in discriminating between classes can be obtained.

**Fig 3.**
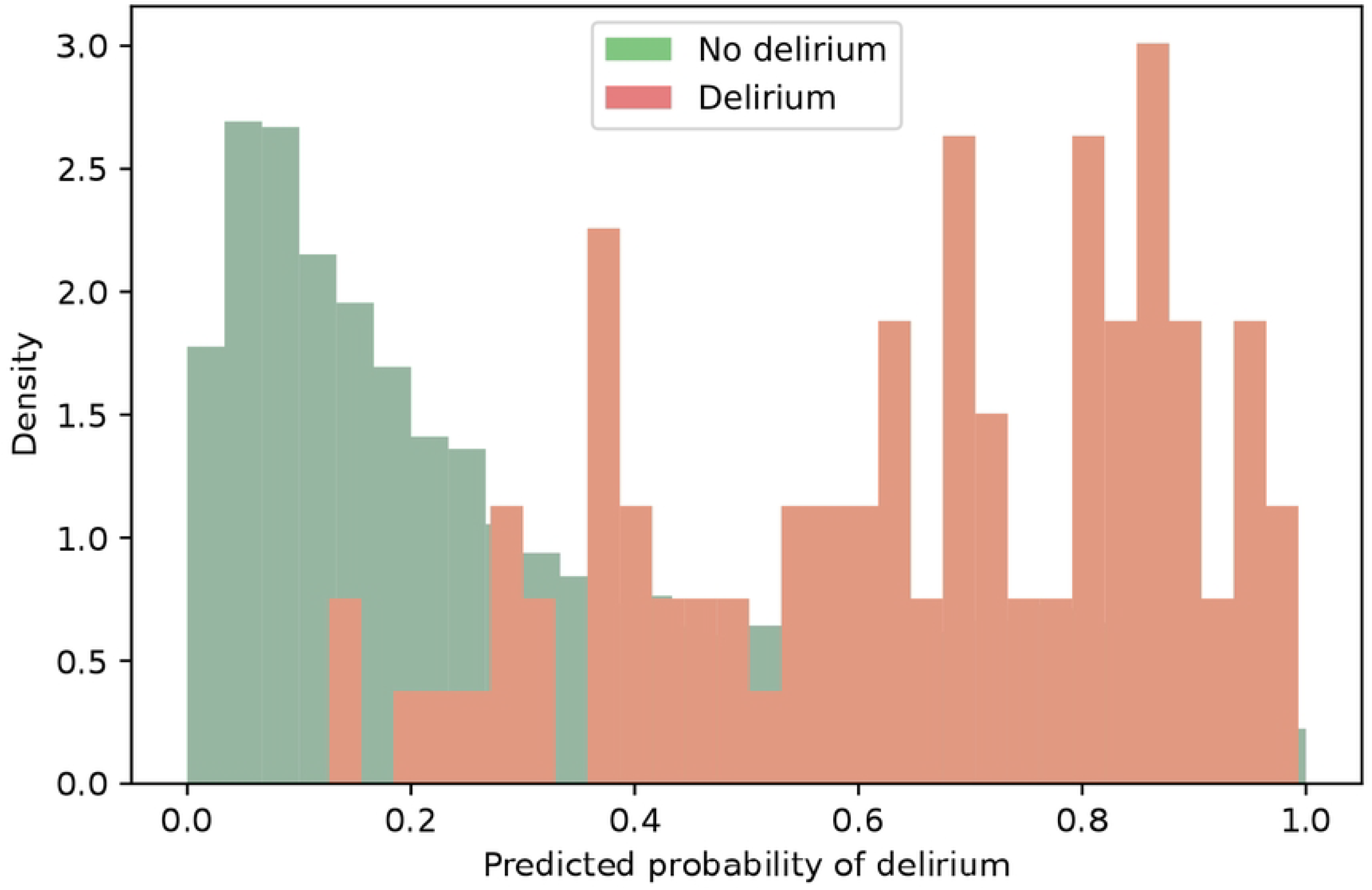
Feature importance for the random forest classifier.

## Discussion

On a large cohort of abdominal surgery patients, we demonstrated that a random forest classifier can be used to predict postoperative delirium with a strong AUC-ROC. Furthermore, this cohort represented a more challenging classification problem as it was significantly imbalanced, with an incidence of approximately 1.15%, owing to the inclusion of adults of any age. We believe such a choice is necessary for a comprehensive POD risk prediction tool, since, although it is more common in elderly adults, POD is a real risk of surgery for any pediatric or adult patient [35, 36]. Furthermore, the clinical validity of the proposed model is corroborated somewhat by the feature importances displayed in Figure 3. We note that comorbidity indices such as the Elixhauser and Charlson comorbidity indices are known to be associated with postoperative delirium across multiple types of surgeries, including abdominal surgeries [22, 51]. Age, another feature prioritized by the random forest classifier, is well-known to be associated with POD [12]. The model also highlights some laboratory markers which are coherent with the presumed pathophysiology of POD. For instance, neuroinflammation is one mechanism which is thought to contribute to POD [3], and an increased minimum preoperative platelet count is a discriminatory factor for the model. Since platelets are an acute phase reactant and are elevated in inflammation [52], this is most likely a surrogate measure for neuroinflammation.

The random forest classifier is able to model more complex patterns than simple classifiers such as logistic regression. However, given the success that neural networks have had in diverse areas of healthcare, and that they are universal function approximators [53], it is noteworthy that they do not perform as strongly as the random forest classifier. It is expected that there is a complex, nonlinear relationship between the independent variables and POD, which, given a sufficient amount of high-quality data, could be effectively modelled by a neural network. Thus, this is possibly due to the relatively small cohort size, compared to other applications of deep learning with millions of observations, or potentially due to the relatively low proportion of positive instances in the dataset. Alternatively, a more complex architecture, rather than a multilayer perceptron, may be able to better model the dataset. Either of these two factors, or a combination, would preclude a neural network from accurately learning a pattern between the input variables and POD.

While we believe that the selection of this cohort was optimal with respect to a broad population and a specific type of surgery, there are some limitations. Most notably, although the cohort itself was rather large, the incidence of POD was lower than expected. Large ranges have been reported across a number of studies, with incidence in elderly patients ranging from 4%–53% [54]. While our focus was not only on the elderly populace, and thus the incidence of POD would be expected to decrease, an incidence of 1.15% is arguably low. This may be due to using ICD-9 and ICD-10 coding for recognizing postoperative delirium, which relies on both the recognition of delirium (which is underdiagnosed [55]) and the entry of the corresponding ICD-9 or ICD-10 codes into the electronic medical record. In light of this, it is necessary to carefully interpret model metrics. In particular, while metrics such as accuracy and precision (also referred to as positive predictive value) vary with prevalence, we have also reported metrics that are more stable with respect to prevalence, such as sensitivity, specificity, and AUC-ROC. One final limitation lies in the manner in which the MIMIC-IV dataset was created. Notably, all data from patients who visited the emergency department or were admitted to the intensive care unit at the Beth Israel Deaconess Medical Hospital between 2008–2022 were extracted from the corresponding medical records for inclusion into the MIMIC-IV dataset. Therefore, although there is no guarantee as to whether these admissions or emergency visits were prior to or after the abdominal surgeries, it is likely that this cohort represents a more ill population than is standard.

Given the nature of the dataset and the distribution of events, future work investigating how our approach and our findings generalize to other datasets would be welcome. In particular, whether or not markers of inflammation remain as valid predictors of POD would be especially clinically useful information. While we believe that a tool to predict POD in a broad population for a specific type of surgery is most clinically relevant, it would also be helpful to verify the incidence of delirium in a cohort constructed in a similar manner. In particular, while we expect that the incidence in this cohort is low, due at least in part to considering all patients instead of just elderly patients, it is unclear to what extent our approach and findings would generalize to other populations owing to the low incidence. Finally, while we highlighted the efficacy of comorbidity indices and laboratory values in the prediction of POD, with larger cohort sizes, an increased number of input variables could be considered, as well as other methods of modelling the data. For instance, given their success in many domains of machine learning, transformers adapted for tabular data [56] could be investigated alongside a number of other architectures.

## Conclusion

In this study, automatic POD prediction was made using common ML models, including random forests, support vector machines, extreme gradient boosted machines, neural networks, and logistic regression. To predict POD, 7215 adult patients with 8026 abdominal surgeries as part of the MIMIC-IV dataset were included in the study. Among the extensive demographic information and commonly measured lab values in this dataset, age, gender, inflammatory markers, comorbidities, inflammatory markers, and alcohol use carried substantial weight in predicting POD. Among all tested ML models, the random forest model outperformed the others in sensitivity (73.11), specificity (71.14), and AUC-ROC (0.800). Summarily, these results implied that the random forest model could be more utilized in the future for automatic POD prediction and highlighted the importance of preoperative inflammatory markers in addition to commonly known risk factors. Also, inasmuch as they are modifiable, risk factors such as comorbidities should be managed to reduce the rate of POD.

While we note that these findings are important, especially for the identification of certain risk factors and with respect to broadening the population for which POD can be predicted, future work could investigate how our approach generalizes to other datasets in different geographical and ethnic settings. Specifically, future studies could validate whether the strong predictive features of POD identified in this study also stand in other various dataset settings. Additionally, further studies investigating whether these or similar results generalize to other cohorts with different incidences of POD would be most welcome.

## Data Availability

Data used in this manuscript is freely available via PhysioNet (https://physionet.org/content/mimiciv/3.1/).

## Notes

### Competing Interest Statement

The authors have declared no competing interest.

### Funding Statement

The author(s) received no specific funding for this work.

### Author Declarations

This study used de-identified data from the MIMIC-IV database, hosted on PhysioNet. The collection of patient information and creation of the MIMIC-IV resource were reviewed and approved by the Institutional Review Boards (IRBs) of the Massachusetts Institute of Technology (MIT, Cambridge, MA, USA) and Beth Israel Deaconess Medical Center (BIDMC, Boston, MA, USA) (MIT IRB \#0403000206 BIDMC IRB \#2001-P-001699/14), who granted a waiver of informed consent and authorized data sharing. All patient data are fully de-identified in accordance with the Health Insurance Portability and Accountability Act (HIPAA) Privacy Rule. Therefore, the use of MIMIC-IV data for research purposes is considered exempt from additional institutional review board approval or informed consent requirements.

